# Geographic distribution of conversion therapy prevalence in Canada

**DOI:** 10.1101/2022.12.27.22283977

**Authors:** Amrit Tiwana, Travis Salway, Julia Schillaci-Ventura, Sarah Watt

**Affiliations:** Faculty of Health Sciences, Simon Fraser University, Burnaby, British Columbia, Canada; Department of Psychology, Simon Fraser University, Burnaby, British Columbia, Canada

## Abstract

Conversion therapy practices (CTPs) are discredited efforts that target lesbian, gay, bisexual, trans, queer, Two-Spirit, or other (LGBTQ2S+) people and seek to change, deny, or discourage their sexual orientation, gender identity, and/or gender expression. This study aims to investigate the prevalence of CTPs across Canadian provinces and territories and identify whether CTP bans are associated with decreasing prevalence. We analyzed 119 CTPs reported from 31 adults (18+) in Canada who have direct experience with CTPs, know people who have gone to CTPs, or know of conversion therapy practitioners by using a 2020 anonymous online survey. Mapping analysis was conducted using ArcGIS Online. CTP prevalence was compared between provinces/territories with and without bans using chi-square tests. We found 3 provinces and 11 municipalities had CTP bans. The prevalence of CTPs in provinces/territories with a ban is 2.34 per 1,000,000 population (95% CI 1.65, 3.31). The prevalence of CTPs in provinces/territories without a ban is 4.13 per 1,000,000 population (95% CI 3.32, 5.14). Accounting for the underlying population, provinces/territories with the highest prevalence of CTPs are New Brunswick (6.69), Nova Scotia (6.50), and Saskatchewan (6.37). Findings suggest only 55% of Canadians are protected under CTP bans. The prevalence of CTPs in provinces/territories without a ban is 1.76 times greater than in provinces/territories with a ban. CTPs are occurring in most provinces/territories, with higher prevalence in the west and the Atlantic. Findings will help inform policymakers and legislators as they are increasingly acknowledging CTPs as a threat to the health and well-being of LGBTQ2S+ people.

## Introduction

“ Conversion therapy” practices (CTPs) are widely discredited, yet they continue to occur across the globe, including Canada [1]. CTPs refer to any practice designed to change, deny, or discourage: one’s feelings of sexual attraction to members of the same gender; lesbian, gay, bisexual, trans, queer, Two-Spirit, or other (LGBTQ2S+) identity; non-conforming gender expression; or gender identity that differs from sex at birth [2-3]. These practices come in many forms and are also sometimes referred to by other names including sexual orientation and gender identity and expression change efforts (SOGIECE), aversion therapy, reparative therapy, and ex-gay ministries [4].

SOGIECE are not monolithic or clearly delineated practices, given that they can happen in many forms. In some contexts, a distinction between CTPs and SOGIECE is useful. CTPs are typically organized (i.e., structured activities) and circumscribed (e.g., a certain number of sessions with a practitioner); these practices are, metaphorically, only the “ tip of the iceberg” [5]. CTPs are underpinned by more prevalent practices including other forms of SOGIECE and cissexist and heterosexist attitudes. SOGIECE have a similar aim as CTPs, but also includes practices that are less well-defined and advertised. For example, a conversation between a parent and a child in which the child is dissuaded from living with or adopting an LGBTQ2S+ identity constitutes as SOGIECE. Overall, both CTPs and SOGIECE are enabled and condoned by widespread heterosexism and cissexism in societies [3].

CTPs are performed by licensed health care providers (e.g., psychologists, psychiatrists, and psychotherapists) and unlicensed practitioners [6]. It has also been performed by ex-gay ministries (e.g., EXODUS) and religious leaders (e.g., pastoral counsellors) [7]. CTP attempts may also involve parents, government agencies, and school personnel.^6^ It may be performed one-on-one in an office or in groups at retreats or conferences [4]. Providers may perform CTPs for money or for free [4]. CTPs can include the use of aversive stimuli, individual talk therapy, participation in activities that are typically gendered by social norms and processes (e.g., sports, hunting, fishing, cooking, playing with dolls, etc.), forced sex, and praying and bible study [6].

### The Harms of Conversion Therapy

There is no credible scientific research that proves CTPs are psychologically safe or effective.^8^ Many medical and human rights associations (e.g., World Health Organization, Canadian Psychological Association, Canadian Psychiatric Association, American Medical Association, American Psychological Association, American Academy of Pediatrics, and Amnesty International) have denounced and discredited the effectiveness of CTPs [3-4,8-9]. However, due to continuing discrimination and societal bias against LGBTQ2S+ people and others with non-heterosexual identities/attractions and non-cisgender identities/expressions, some practitioners continue to provide CTPs, particularly to minors, making them vulnerable to harms associated with these practices [9].

Previous research suggests that CTPs negatively impact and stigmatize LGBTQ2S+ people, and can lead to increased anxiety, depression, self-hatred, post-traumatic stress disorder (PTSD), and many lifelong psychological and social issues [4,8-10]. According to the Sex Now 2011/12 survey, exposure to SOGIECE among Canadian gay, bisexual, and other sexual minority men were positively associated with loneliness, regular illicit drug use, suicidal ideation, and suicide attempt [10].

LGBTQ2S+ health disparities are often conceptualized through the minority stress framework, in which mental health problems found among LGBTQ2S+ people are the result of chronic stressors stemming from the marginalized social status of these individuals, rather than a function of their identity itself [11-13]. This framework offers some insight into the mechanisms through which CTPs may exact harm upon people who experience them [14]. Minority stressors include prejudicial events and conditions that are expressed both interpersonally (e.g., violent attacks and discrimination) and structurally (e.g., laws allowing the rejection of LGBTQ2S+ people in employment) [15]. SOGIECE can also be conceptualized as a minority stressor because it promotes cisheteronormativity as the only acceptable way of life and reinforces the rejection of LGBTQ2S+ identities [15]. Cisheteronormativity refers to “ cissexism and heterosexism which assume cisgender gender identities and heterosexual orientation are more natural and legitimate than those of LGBTQ2S+ people” [5]. As these stressors (including SOGIECE) accumulate, they create emotional (e.g., ruminating on negative messages), cognitive (e.g., feeling negative about oneself and/or hopeless about one’s future), and social (e.g., social anxiety/avoidance) maladaptive responses, eventually leading to diagnosable conditions like anxiety or depression [14].

### Prevalence of Conversion Therapy

According to the survey results from Sex Now 2019 (SN2019), an estimated 50,000 Canadian gay, bisexual, and other sexual minority men have attended CTPs at some point in their lives [10]. This corresponds to 1 in 10 gay, bisexual, and other sexual minority men (10%) having reported experiencing CTPs. In addition, the previous Trans PULSE Canada community-based survey found that 11% of transgender and non-binary people had undergone SOGIECE [16]. Additionally, results from SN2019 suggest that transgender and non-binary people experience a higher burden of exposure to CTPs than cisgender people [17].

Results from SN2019 did not find a difference in the prevalence of CTPs across provinces and territories in Canada [10]. However, it did find uneven exposure of CTPs across groups defined by age, gender identity, immigration, and ethnicity [17]. CTPs were even more common (i.e., more than 10%) among subgroups including youth 15-19 years of age (13%), immigrants (15%), and racial/ethnic minorities (11-22%) [17]. Besides the annual Sex Now Surveys and Trans PULSE Canada Survey, no data are available regarding the distribution of CTPs in Canada. Further, no data are available examining the geospatial patterning of CTPs across and within different geographic regions of Canada, highlighting the importance of the current study.

### Attempts to Ban Conversion Therapy

As of November 11, 2021, five Canadian provinces and territories (Ontario, Nova Scotia, Prince Edward Island (PEI), Quebec, and Yukon) and dozens of Canadian municipalities (including Calgary, Edmonton, and Vancouver) have enacted legislative bans on CTPs [18]. On December 4, 2021, the Canada House of Commons voted unanimously to pass Bill C-4, *an Act to Amend the Criminal Code (conversion therapy)*. Bill C-4 subsequently was passed by the Senate and received Royal Assent on December 8, 2021. Bill C-4 includes provisions to prohibit causing a person to undergo CTP for, removing a child from Canada to undergo CTP outside Canada, and advertising or financially benefiting from CTP. This legislation represents an important milestone in the government’s commitment to protecting LGBTQ2S+ people. As policymakers and legislators are increasingly acknowledging CTPs as a threat to the health and well-being of LGBTQ2S+ people, up-to-date data on the geospatial patterns of CTP prevalence are urgently needed.

Given that CTPs are widely discredited and denounced practices, many conversion therapy providers do not advertise themselves as “ conversion therapists” [19]. Wide denunciation and steps towards criminalization may have led conversion therapy providers to operate ‘underground’ and advertise their services in covert ways [20]. These providers may not refer to their services as “ conversion therapy” and may instead use vague terms to allude to their approach to diverse sexual orientations or gender identities (e.g., healing “ sexual brokenness”) [20]. For these reasons, it is challenging to track down the locations where CTPs continue to operate. Establishing methods and data sources that enable ongoing monitoring of CTPs is critical in this context.

### The Purpose of this Research

For this study, data were collected from a small national sample of Canadians who have direct experience with CTPs, know people who have gone to CTPs, or know of conversion therapy practitioners in 2020. The goal of the survey was to better understand the nature and scope of SOGIECE, or CTPs in Canada. The primary objective of this thesis was to estimate the spatial patterning of prevalent CTPs in Canada.

## Materials and Methods

### Participants

An anonymous online survey using SurveyMonkey was conducted between August 18 and December 2, 2020. The survey was conducted in French and English. The content of the questionnaire was developed with the help of conversion therapy survivors and people working to prevent CTPs or reduce the harm associated with CTPs in Canada. To be eligible for the survey, participants must have: experienced CTPs, nearly experienced CTPs, or know someone who has experienced CTPs; been 19 years of age or older; and resided in Canada. The protocol for this study was approved by the Simon Fraser University Research Ethics Board (study 2019s0394).

Participants were recruited using various strategies including word-of-mouth (37% of respondents reported hearing about the survey in this manner), Twitter or Facebook (35%), LGBTQ2S+ community organizations (15%), and participation in an in-depth interview with the research team [21] during January-July 2020 (13%). A total of N = 108 individuals entered the study, of which 19 did not consent to participate and/or were ineligible to participate. In total, N = 89 completed the entire survey (i.e., responded to the last question). For the present study, a subset of N = 31 (29%) of those who answered the questions regarding known conversion therapy providers in Canada were used in the analysis.

### Measures

Participants were directed to a SurveyMonkey link where they were presented with an informed consent page regarding the study. Specifically, participants were informed about the project, who was conducting and funding the survey, the risks, and benefits of the survey, as well as the eligibility criteria and consent to participate. All participants who completed the survey (N = 89) provided informed consent before completing the questionnaire. Participants were informed they could skip any of the questions they did not wish to answer.

The survey consisted of six parts: (1) recruitment and category of experience with conversion therapy; (2a) details about direct SOGIECE experience; (2b) details about knowledge of SOGIECE; (3) knowledge of conversion therapy practitioners; (4) legislative action against conversion therapy; and (5) social-demographic characteristics. To determine eligibility, participants were asked: “ How would you describe your experience with conversion therapy (i.e., structured activity to deny or suppress your LGBTQ2 identity)? Please check all that apply.” Respondents were directed to part (3) of the survey, “ knowledge of conversion therapy practitioners”, if they selected “ I know of conversion therapy practitioners in Canada” or “ I know that conversion therapy is happening in Canada”. They were then asked to list municipalities, provinces, and territories where these CTPs had taken place, and identify whether the practice was ongoing or historical (i.e., happened in the past).

At the end of the study, all participants were asked to provide social demographic information including their age, place of residence, and racial/ethnic group. Finally, participants were presented with a debriefing letter which included an option to input their email address to learn more about SOGIECE and CTPs, and a list of LBTQ2S+-affirming mental health supports for any participants who may have experienced distress because of the recalling of traumatic experience(s).

### Analyses

To analyze the spatial patterning, maps were created using ArcGIS Online. Participants who answered part (3) of the survey, ‘knowledge of conversion therapy practitioners’ (N = 31) were used in the mapping analysis. The dataset contained a total of 127 reports of 119 ongoing and 8 historical CTPs. Because names/addresses of CTPs were not collected, it was not possible to determine whether the 127 reports are mutually exclusive. Municipal and provincial/territorial spatial units were used in the analysis. Primary analysis focused only on ongoing reports, however additional maps were created combining ongoing and historical reports (see Appendix A and B). Thus, the primary analysis dataset consisted of 119 rows (i.e., observations) and 2 columns (reported municipalities and provinces/territories). These 119 CTPs were identified in 53 different municipalities (with 1-7 CTPs per municipality) and 8 different provinces/territories (with 1-28 CTPs per province/territory). 20 out of the 119 reports identified CTPs in provinces/territories but did not specify a municipality.

Three maps were created: (1) municipal, provincial and territorial CTP bans in Canada, (2) a heat map of ongoing CTPs (using municipal spatial unit), and (3) a choropleth map of CTP prevalence by province/territory (calculated as reports of ongoing CTPs divided by 1,000,000 population per province/territory). Provinces/territories with numerators less than 5 were not interpreted due to statistical instability. As of November 2021, data on current legislative bans were obtained from the Legislation Map on the No Conversion Canada website.^18^ The “ heat map” function in ArcGIS Online was applied to identify hot spots of CTPs across the nation. The map of the bans and the heat map were compared to assess the association between ongoing CTPs and existing bans. We also calculated the prevalence of CTPs in provinces/territories with bans and the prevalence of CTPs in provinces/territories without bans (as of the time of the survey, August 2020) using chi-square tests, to assess differences in prevalence between these two sets of jurisdictions. 95% confidence intervals were added to these measures. The 2016 Census cartographic boundary file by Statistics Canada was used to estimate the population per municipality, province, and territory for the choropleth map [22].

## Results

The age of respondents ranged from 18 to 65+ with 70% of participants under the age of 45. The majority of the respondents resided in the most populous Canadian provinces, i.e., British Columbia, Ontario, and Alberta. The majority of the sample identified with a white ethnic/racial group (93.33%), followed by East Asian, Black, and mixed ethnic/racial groups. A plurality of respondents identified as cis men (38.7%) and gay (41.9%). However, reported gender identity and sexual orientation were diverse, as shown in Table 1.

**Table 1.** Demographic characteristics of participants who report knowing ongoing or historical conversion therapy practices in Canada, (N = 31).

| **Demographic Characteristics** |  | **Total**<br>**n (%)** |
| --- | --- | --- |
| **Age**<br>**(years)** | 18-24<br>25-34<br>35-44<br>45-54<br>55-64<br>65+<br>N/A | 6 (20)<br>9 (30)<br>7 (22.6)<br>3 (10)<br>2 (6.7)<br>1 (3.3)<br>3 (10) |
| **Area of Residence**<br>**(Province/Territory)** | British Columbia<br>Alberta<br>Saskatchewan<br>Manitoba<br>Ontario<br>Quebec<br>Atlantic<br>Territories<br>N/A | 8 (26.7)<br>4 (13.3)<br>0<br>0<br>9 (30)<br>3 (10)<br>3 (10)<br>0<br>3 (10) |
| **Race/Ethnicity** | White<br>Black<br>East Asian<br>Indigenous<br>Mixed<br>N/A | 19 (63.3)<br>2 (6.7)<br>3 (10)<br>1 (3.3)<br>2 (6.7)<br>3 (10) |
| **Gender Identity\*** | Cis Men<br>Cis Women<br>Trans Men<br>Trans Women<br>Non-Binary<br>Genderqueer<br>Genderfluid | 12 (38.7)<br>5 (16.1)<br>5 (16.1)<br>2 (6.5)<br>5 (16.1)<br>3 (9.7)<br>2 (6.5) |
| **Sexual Orientation\*** | Gay<br>Queer<br>Lesbian<br>Bisexual<br>Pansexual<br>Asexual<br>Heterosexual/straight<br>Queer Stone Butch | 13 (41.9)<br>12 (38.7)<br>6 (19.4)<br>4 (12.9)<br>4 (12.9)<br>2 (6.5)<br>4 (12.9)<br>1 (3.2) |

*Note.* N/A refers to “not available” meaning the respondent did not fill out the demographic characteristic of interest. \*Participants are permitted to select more than one option.

Data collection took place between August and December 2020, therefore legislative bans passed on CTPs after August 2020 are not considered in the results. Then, only three provinces (Ontario, PEI, and Nova Scotia) and eleven municipalities had legislative bans in place. 19,459,172 out of 35,151,728 Canadians were protected under legislative bans as of August 2020, which corresponds to 55.1% of the total Canadian population (Table 2). Fig 1 is a map visualizing the bans passed as of August 2020.

**Table 2.** List of the dates of legislative bans passed on conversion therapy practices (CTPs) in Canadian municipalities, provinces, and territories.

| Municipality | Province/Territory | Date of Legislative Bans on CTPs | Canadian Population, 2016 | Cumulative Percentage of Canadian Population Protected Under Legislative Ban |
| --- | --- | --- | --- | --- |
|  | Ontario | 2015 | 13,448,494 | 38.3% |
| Vancouver | British Columbia | June 2018 | 631,486 | 40.1% |
|  | Nova Scotia | 2018 | 923,598 | 42.7% |
| Montreal | Quebec | August 2019 | 1,704,694 | 47.5% |
| Strathcona County | Alberta | September 2019 | 98,044 | 47.8% |
| Edmonton | Alberta | December 2019 | 932,546 | 50.5% |
| St. Albert | Alberta | December 2019 | 65,589 | 50.7% |
|  | Prince Edward Island | 2019 | 142,907 | 51.1% |
| Wood Buffalo (Fort McMurray) | Alberta | January 2020 | 71,589 | 51.3% |
| Saint John | New Brunswick | January 2020 | 67,575 | 51.5% |
| Rocky Mountain House | Alberta | February 2020 | 6,635 | 51.5% |
| Calgary | Alberta | March 2020 | 1,239,220 | 51.5% |
| Spruce Grove | Alberta | April 2020 | 34,066 | 55.0% |
| Lethbridge | Alberta | July 2020 | 92,729 | 55.1% |
| **Data collection occurred August-December 2020. Therefore, the following legislative bans are not accounted for in the analysis.** |  |  |  |  |
| Beaumont | Alberta | November 2020 | 17,396 | 55.4% |
|  | Yukon | November 2020 | 35,874 | 55.4% |
|  | Quebec | December 2020 | 8,164,361 | 78.1% |
| Kingston | Ontario | January 2021 | 123,798 | 79.1% |
| Saskatoon | Saskatchewan | February 2021 | 246,376 | 79.8% |
| Nanaimo | British Columbia | March 2021 | 90,504 | 80.0% |
| Fort Saskatchewan | Alberta | July 2021 | 24,149 | 80.1% |
| Strathmore | Alberta | July 2021 | 13,756 | 80.2% |
| Thunder Bay | Ontario | July 2021 | 107,909 | 80.5% |
| Regina | Saskatchewan | August 2021 | 215,106 | 81.1% |

*Note.* Data collection started in August 2020 and ended in December 2020. Legislative bans passed after August 2020 are not accounted for in the analysis.

**Fig 1.**
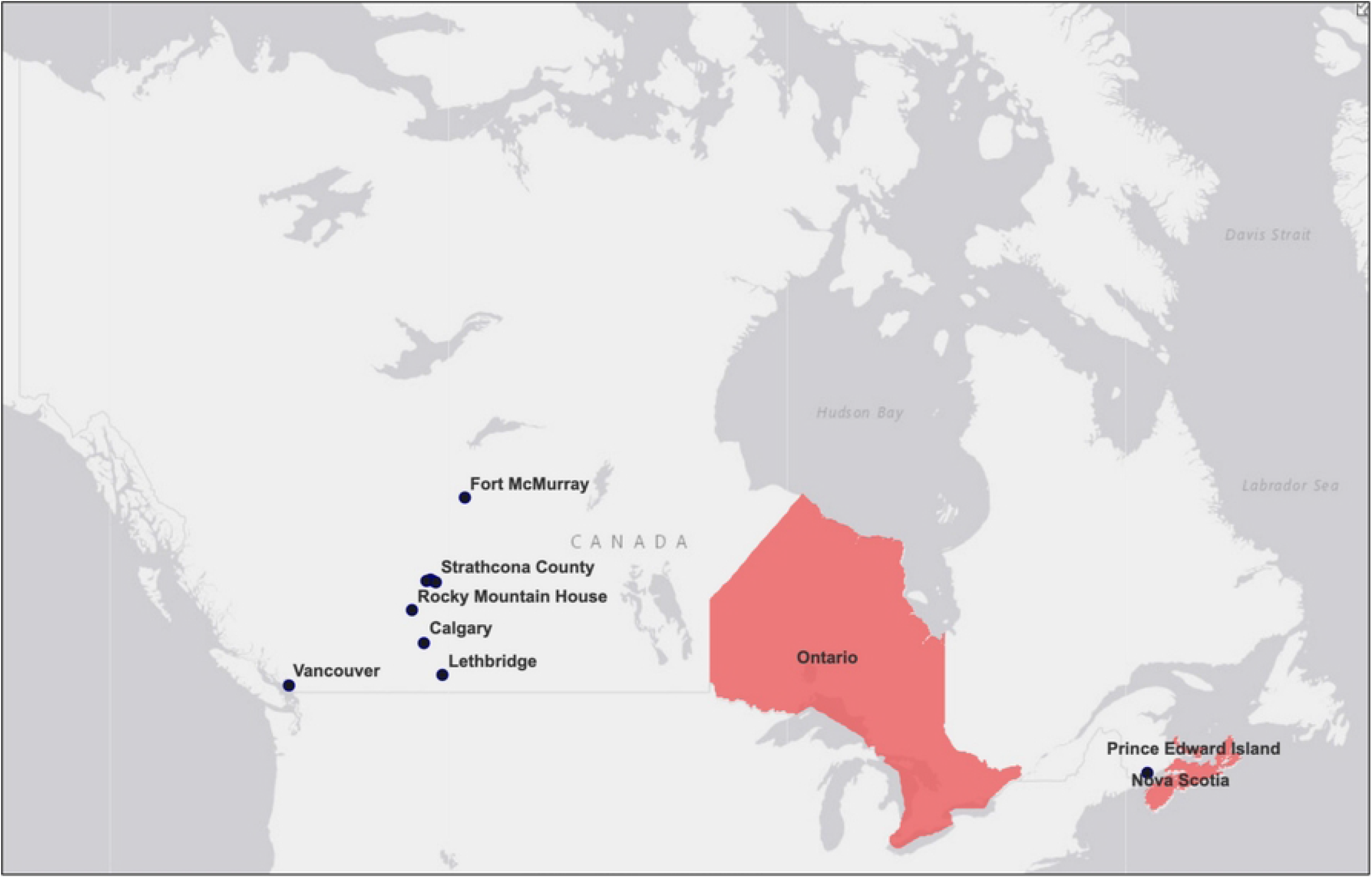
Municipal, provincial, and territorial legislative bans on conversion therapy practices (CTPs) in Canada as of August 2020.

31 out of the 89 participants who answered part (3) of the survey, ‘knowledge of conversion therapy practitioners’ reported 119 CTPs (reports per participant ranged from 1-31, mean of 4). 20 (16.8%) locations were reported at the provincial/territorial level and 99 (83.2%) locations were reported at the municipal level. The prevalence of ongoing CTPs in provinces and territories with bans as of August 2020 is 2.34 per 1,000,000 population (95% CI 1.65, 3.31) (Table 3). The prevalence of ongoing CTPs in provinces and territories without bans as of August 2020 is 4.13 per 1,000,000 population (95% CI 3.32, 5.14). No data were reported regarding CTPs in the Northwest Territories and Nunavut and therefore are not included in the calculations.

**Table 3.** Prevalence of ongoing conversion therapy practices (CTPs) in Canadian provinces and territories with bans versus without bans at the provincial level.

|  | Number of practices identified (numerator) | Prevalence (CTPs per 1,000,000) (95% confidence interval) |
| --- | --- | --- |
| Ongoing CTPs per 1,000,000 population in provinces/territories <u>WITH</u> bans (as of August 2020): | 34 | 2.34<br>(1.65, 3.31) |
| Ongoing CTPs per 1,000,000 population in provinces/territories <u>WITHOUT</u> bans (as of August 2020): | 85 | 4.13<br>(3.32, 5.14) |

*Note.* No data were reported regarding CTPs in the Northwest Territories and Nunavut. Therefore, Nunavut and the Northwest Territories are not included in the calculations above.

A heat map was created to examine a more granular prevalence of reported ongoing CTPs in Canada using count data without adjustment of the underlying population. We used the municipal spatial unit for the heat map because data containing just the reported province/territory were misleading. As shown in Fig 2, ongoing CTPs were identified across the nation. The majority of the cases and hotspots are clustered around the South of Canada. This is no surprise given that two-thirds (66%) of the Canadian population lives within 100 kilometres of the southern Canada-United States (US) border, an area that represents approximately 4% of Canadian land [23].

**Fig 2.**
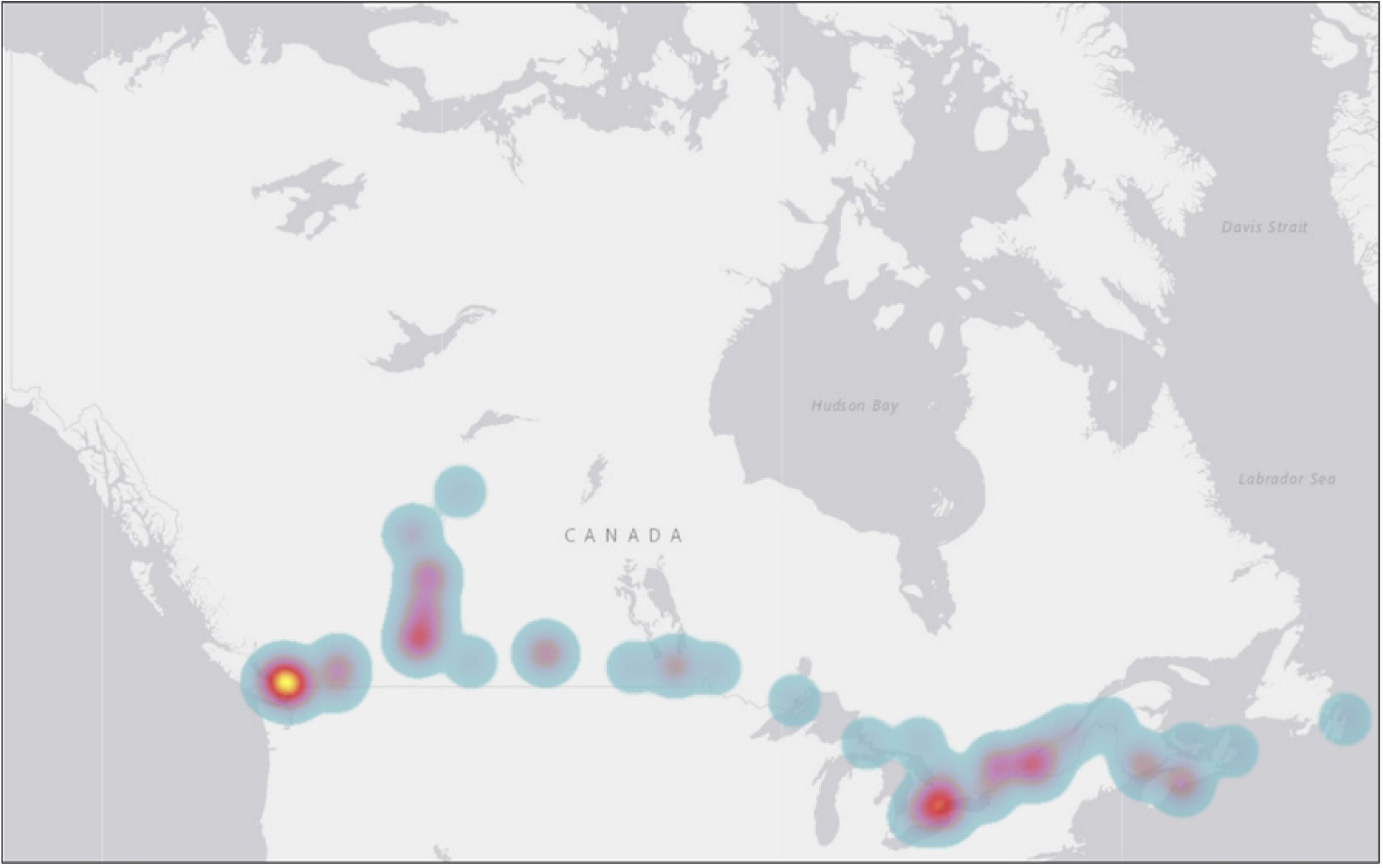
A heat map of the prevalence of reported ongoing conversion therapy practices (CTPs) in Canada using count data (N = 99) *Note*. No data were reported regarding the prevalence of CTPs in the Northwest Territories and Nunavut.

Fig 3 displays the prevalence of CTPs divided by 1,000,000 population per province/territory. When accounting for the underlying population, the provinces and territories with the highest prevalence of reported ongoing CTPs are New Brunswick (6.69), Nova Scotia (6.50), and Saskatchewan (6.37). Provinces and territories with moderate prevalence include British Columbia (6.08), Alberta (5.90), and Manitoba (3.91), Provinces with lower prevalence include Ontario (2.01), and Quebec (1.59). The following provinces/territories had <5 reports of conversion practices and therefore are not further interpreted: Yukon (55.75), PEI (7.00), and Newfoundland and Labrador (1.92). No data were reported regarding the prevalence of ongoing CTPs in the Northwest Territories and Nunavut.

**Fig 3.**
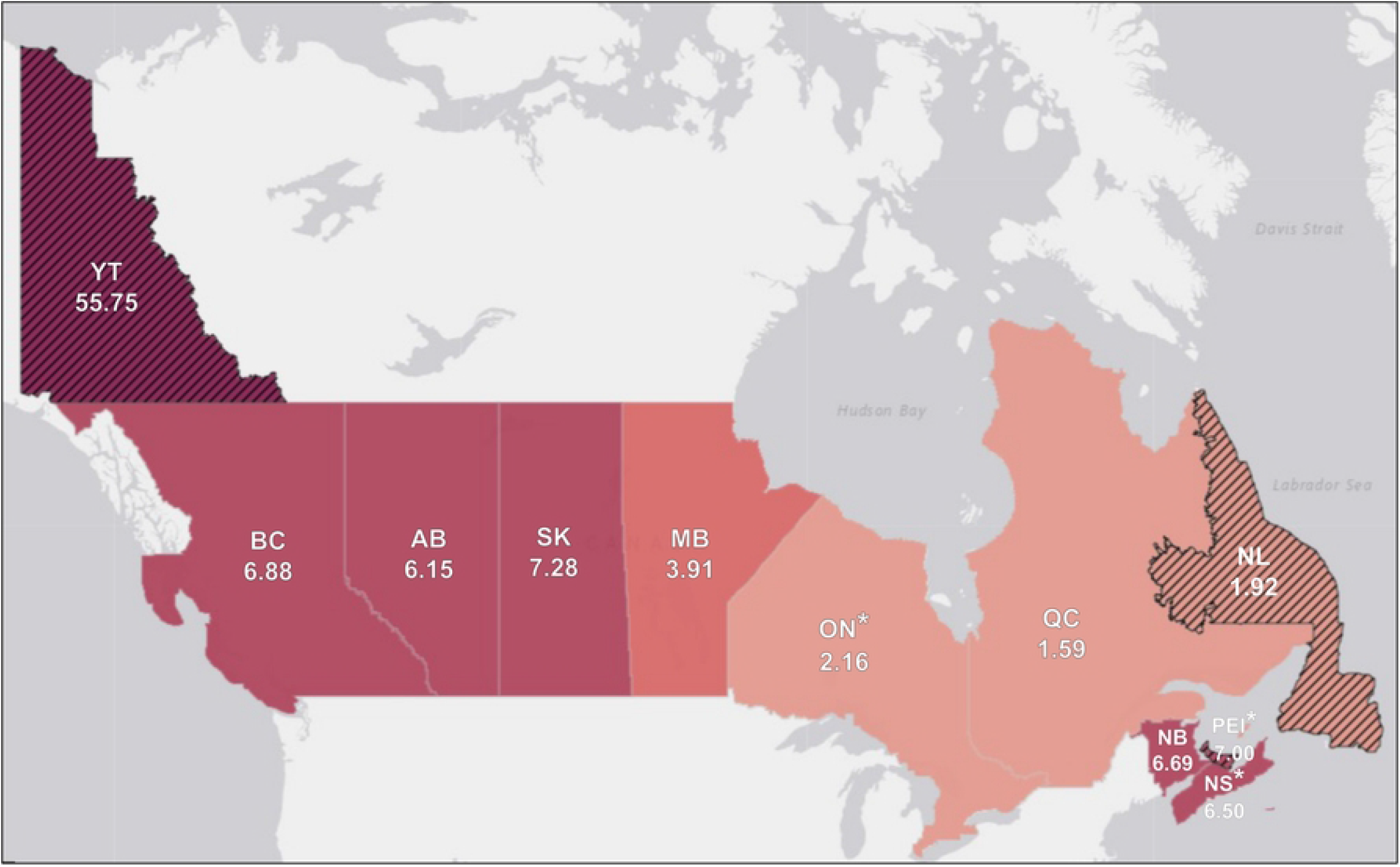
Prevalence of ongoing reported conversion therapy practices (CTPs) divided by 1,000,000 population per province/territory (N = 119) *Note*. *Province/territory has implemented a ban as of August 2020. Please exercise caution in interpreting estimates from the following provinces/territories with numerators <5: Yukon (YT) (55.75), Prince Edward Island (PEI) (7.00), Newfoundland and Labrador (NL) (1.92). No data were reported regarding the prevalence of CTPs in the Northwest Territories and Nunavut.

## Discussion

This non-probabilistic survey of adults in Canada with direct or indirect experiences of CTPs provides early insights on the spatial patterning of the prevalence of CTPs in Canada. Findings suggest only 19,459,172 out of 35,151,728 Canadians were protected under the legislation on CTPs at the time of the study. This number corresponds to 55.1% of the total Canadian population protected. Further, CTPs are occurring in the majority of Canadian provinces and territories, with higher prevalence in the west and the Atlantic.

These findings are similar to the results of a study that examined exposure to psychological attempts to change a person’s gender identity from transgender to cisgender (PACGI) among transgender people in the US, lifetime and between the years 2010 and 2015, by US state [24]. The findings from this study suggested that practices continue to occur in every US state as recently as the period 2010 to 2015, despite major US medical organizations identifying PACGI as ineffective [24] and legislative bans passed in 20 states and more than 100 municipalities in the US [25]. As with the Canadian map (Fig 3), The PACGI study found an elevated prevalence of PACGI in western jurisdictions of the US.

The prevalence of CTP in provinces/territories without legislative bans as of August 2020 was approximately 1.76 times greater than the prevalence in provinces/territories with bans (Ontario, PEI, Nova Scotia). While this is encouraging, there are several reasons to be cautious in interpreting this finding. The data used in this analysis does not allow us to control for other factors happening within the provinces/territories with bans that have deterred CTPs (i.e., confounding). For example, we did not account for whether there might have been an effect of municipal bans. Additionally, we cannot know from these data whether bans are enforced. We cannot say whether legislative bans caused CTPs to shut down because we do not know whether these services started or stopped practicing before or after the bans. In other words, we cannot assess the temporal relationship between time and practices taking place. For these reasons, the association between the lower prevalence found in provinces/territories with bans and higher prevalence found in provinces/territories without bans may not be real.

### Limitations

First, the sample size was small. The prevalence calculated may be unstable because it is based upon only a small number of reports extrapolated over a large population [26]. Second, the sample largely came from LGBTQ2S+ community organizations or conversion therapy survivors and thus constitutes a non-probabilistic subset of the total target population (e.g., all people in Canada). Evidence shows that non-probability surveys tend to overrepresent employed, high-income-earning, and gay-identified sexual minorities [27]. Third, since the survey was developed in Vancouver, British Columbia, it is likely more CTPs were reported in this region compared to other parts of Canada, which is reflected in the geographic patterning of the data. Fourth, the language of the item used to measure conversion therapy may not encompass people’s experiences. For example, people may interpret their experience as more of a form of SOGIECE rather than conversion therapy and thus may choose to not report the practice. Lastly, we cannot assess legislation impacts and whether it reduced the prevalence of CTPs because we do not know if the identified practice took place before or after legislation was passed.

### Future Research

Given that CTPs persist across Canada, we recommend that the following actions be taken in collaboration with conversion therapy survivors, community organizations and multiple levels of government. The results of this study should be confirmed and repeated with a larger sample size to ensure underscore representativeness. One idea to deal with the small sample size problem is to bring additional years of data into the analysis to increase the size of the numerator [26]. In addition to this, recruitment methods should be expanded to make interpretations more generalizable to the Canadian population. There must be continued efforts to monitor how conversion therapy practitioners continue to operate, even in places where bans have been implemented. Monitoring where CTPs continue to occur will help keep legislators accountable and identify where supportive LGBTQ2S+-affirming environments are most needed.

## Conclusion

The ongoing occurrence of CTPs is a serious public health issue impacting the health and well-being of thousands of LGBTQ2S+ Canadians. Findings from this study should be taken into consideration as Bill C-4 is implemented and enforced in Canada. Combined with legislative ban efforts, the Canadian government should work to deter SOGIECE and CTPs, while encouraging LGBTQ2S+-affirming environments by supporting gay-straight alliances, pride flags, and other interventions that remind LGBTQS2+ people that their identities are valid [3]. Finally, education is required to reduce the prejudice of LGBTQ2S+ people to ultimately put an end to CTPs.

## Data Availability

Data cannot be shared publicly because it contains sensitive geographic data. Data are available from the Simon Fraser University Vault and Simon Fraser Research Ethics Board (contact via) for researchers who meet the criteria for access to confidential data.

## Acknowledgments

My research team is grateful to the following individuals who provided insight and feedback on the questionnaire drafts and/or who helped to promote the survey: A.J. Lowik, Beth Carlson-Malena, David Kinitz, Elisabeth Dromer, Geron Malbas, Keith Murray, Kiffer Card, Michael Kwag, Nicholas Schiavo, and Trevor Goodyear.

## Supporting Information

**S1 Fig. Sensitivity analysis: heat map of the prevalence of reported ongoing and historical conversion therapy practices (CTPs) in Canada using Count Data (N = 106)**

*Note*. No data were reported regarding the prevalence of CTPs in the Northwest Territories and Nunavut.

**S2 Fig. Sensitivity analysis: prevalence of ongoing and historical reported conversion therapy practices (CTPs) divided by 1**,**000**,**000 population per province/territory (N = 127)**

*Note*. *Province/territory has implemented a ban as of August 2020. Please exercise caution in interpreting estimates from the following provinces/territories with numerators <5: Newfoundland and Labrador (1.92), Prince Edward Island (7.00), and Yukon (55.75). No data were reported regarding the prevalence of CTPs in the Northwest Territories and Nunavut.

## Notes

### Competing Interest Statement

The authors have declared no competing interest.

### Funding Statement

This study was supported by the Canadian Institutes of Health Research (FRN: PCS - 168193), Andrew Beckerman, and the Victoria Foundation. The funding sources did not play a role in the study design, the collection and analysis of data, the decision to publish or the preparation of the manuscript.

### Author Declarations

The protocol for this study was approved by the Simon Fraser University Research Ethics Board (study 2019s0394).

